# Associations between natural sunlight exposure and brain structural markers: a prospective study in the UK Biobank

**DOI:** 10.1101/2023.10.12.23296944

**Authors:** Huihui Li, Fusheng Cui, Tong Wang, Weijing Wang, Dongfeng Zhang

## Abstract

**Objective:** Sunlight is closely intertwined with daily life. It remains unclear whether there are associations between sunlight exposure and brain structural markers.

**Methods:** This longitudinal study utilized baseline data (2006-2010) and follow-up data (2014+) from the UK Biobank. General linear regression analysis was employed to compare the differences in brain structural markers among different sunlight exposure time groups. Stratification analyses were performed based on sex, age, and diseases (hypertension, stroke, diabetes). Limiting cubic splines were performed to examine the dose-response relationship between natural sunlight exposure and brain structural markers, with further stratification by season. To control environmental and genetic factor, we adjusted PM2.5 and PRS for Alzheimer’s disease.

**Results:** A total of 27,474 participants were included in the final analyses. The association of sunlight exposure time with brain structural markers was found in the upper quartile compared to the lower quartile. Prolonged natural sunlight exposure was associated with the volumes of total brain (β: -0.051, P < 0.001), white matter (β: -0.031, P = 0.023), gray matter (β: -0.067, P < 0.001), and white matter hyperintensities (β: 0.059, P < 0.001). These associations were more pronounced in males and individuals under the age of 60. With daily sunlight exposure approximately exceeding 2 hours, we observed that total brain volume and gray matter volume decreased, while white matter high hyperintensity volume increased with prolonged sunlight exposure duration.

**Conclusions:** This study demonstrates that prolonged exposure to natural sunlight is associated with brain structural markers change. These findings offer new insights into the mechanisms underlying the association between natural sunlight and brain health.

## Introduction

Sunlight is closely associated with human health. Sunlight plays a crucial role in maintaining overall health by participating in multiple processes such as skin synthesis of vitamin D[1, 2] and regulating the circadian rhythm[3, 4]. However, inappropriate exposure to ultraviolet (UV) radiation from sunlight can result in both acute and chronic health consequences, including skin cancer,[5] sunburn (erythema),[6] immunosuppression,[7] DNA damage,[8] and more. The UV radiation has the potential to suppress cell-mediated immune function, leading to inflammatory responses,[7] while the inflammatory responses are recognized as one of the risk factors for dementia.[9] Additionally, worsening air pollution has contributed to the thinning of the ozone layer, reducing its capacity to absorb UV radiation, which may result in increased UV exposure for individuals.[10] Besides, it has been shown that individuals with lighter skin tones are more susceptible to the effects of UV radiation.[11]

The brain can also be affected by sunlight. The brain volume is primarily composed of white matter and gray matter. White matter occupies more than half of the total human brain volume and is primarily composed of myelinated axons.[12] White matter plays a crucial role in coordinating information transmission and integration among different brain regions.[13, 14] The central nervous system comprises another crucial component known as gray matter, which consists of neurons, glial cells, and microvasculature. These neurons are responsible for processing and transmitting information, while glial cells provide support and protection. The microvascular system supplies oxygen and nutrients to meet the needs of neurons. Brain function relies on the delivery of oxygen and nutrients through blood circulation and depends on the brain’s ability to maintain thermal balance. When exposed to sunlight, more blood flows away from the brain to regulate brain temperature, resulting in a reduced blood flow to the brain, which may lead to brain damage.[15, 16] Experimental studies have found that direct exposure of the head and neck to sunlight radiation can result in a core temperature increase of 1°C, and may impair motor-cognitive functions.[17]

While previous researches have explored the association between sunlight and cognitive function, most studies have primarily focused on the relationship between sunlight and global cognitive function or the occurrence of dementia.[2, 18–21] There remains a gap in the investigation of the associations between natural sunlight exposure and brain structure. Research indicates that changes in brain morphology, such as white matter integrity, may precede and potentially lead to declines in cognitive function,[22] and individual differences in cognitive function are partially explained by variations in brain structure[23]. White matter hyperintensity, as one of the brain structural markers, is associated with pathologies of Alzheimer’s disease.[24, 25] Therefore, natural sunlight exposure may be associated with brain structure.

We used the data from the UK Biobank cohort to address these gaps. The aim of this study is to explore the relationships between sunlight exposure and brain structural markers. Furthermore, since the season, sex, and age differences in the association between sunlight exposure and cognition[2], we further conducted stratified analyses based on these factors to investigate the sunlight-brain structure associations separately. In addition, considering that hypertension,[26, 27] stroke,[28, 29] and diabetes[30, 31] are closely associated with brain structure as well as cognitive impairment, we also tried to analyze the relationships between sunlight exposure and brain structure in these diseases groups, respectively.

## Methods

### Data Sources and Study Design

The UK Biobank is a population-based, large-scale prospective cohort study that recruited over 500,000 participants nationwide from March 2006 to December 2010. After signing the written informed consent forms, all participants completed baseline assessments at one of the 22 assessment centers, which were in England, Scotland, or Wales. These assessments included touchscreen questionnaires, verbal interviews, physical examinations, and the collection of biological samples. Starting in 2014, a subset of participants was invited to four assessment centers for cognitive function questionnaires, imaging scans, and more. The UK Biobank has obtained approval from the Northwest Multi-Center Research Ethics Committee (reference 06/MRE08/65). The specific selection process flowchart is presented in Figure 1.

**Figure 1.**
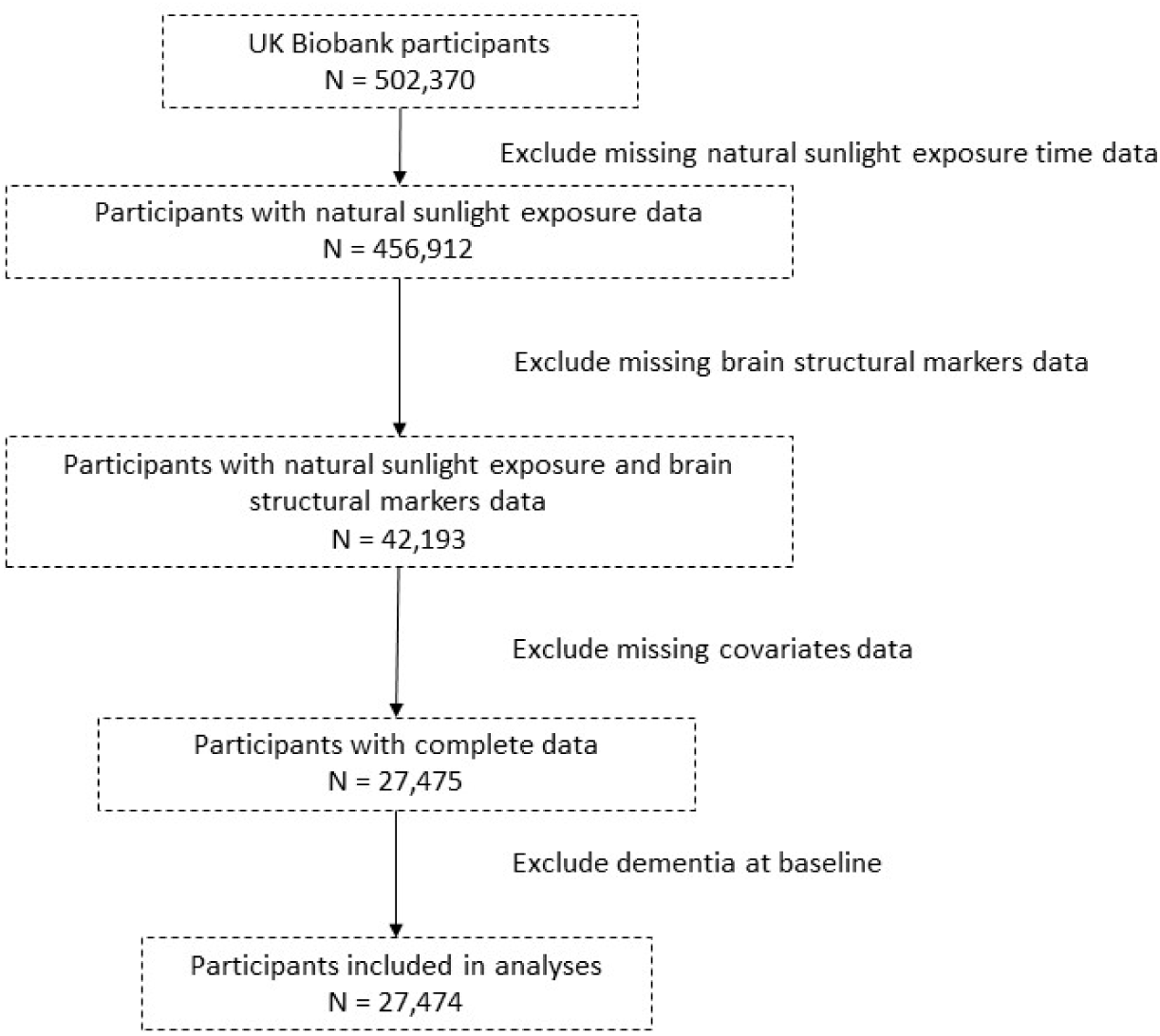
Flowchart illustrating criteria for selection of samples as well as the four analyses performed in the current study.

### Natural Sunlight Exposure Time Measurements

The time spend in summer and winter is collected through touchscreen questionnaires during participants’ visits to the assessment center. Responses of “Don’t know” and “Prefer not to answer” are excluded, and “Less than 1 hour” was redefined as 0 hour. Participants who reported the time exceeding 16 hours in summer and 8 hours in winter were removed based on the effective daylight hours in the UK. The exposure variable was the annual average sunlight exposure time, which was calculated by taking the average outdoor time during both the summer and winter.

### Brain Structural Markers Measurements

The brain structural markers include the volumes of total brain, white matter, gray matter, and white matter hyperintensities. We performed Z-transformations on the brain structural markers based on the mean and standard deviation. T1-weighted data was acquired on a 3T Siemens Skyra scanner using a standard 32-channel head coil. The parameters for the magnetization-prepared rapid gradient-echo imaging sequence were set as follows: resolution: 1×1×1 mm, feld-of-view (FOV): 208×256×256 matrix, duration: 5 min. Subcortical structures were segmented using FIRST (version 5.0), an integrated registration and segmentation tool within FMRIB. Cortical tissue-type segmentation was completed using FAST, FMRIB’s automated segmentation tool. The white matter hyperintensities were calculated based on T1 and T2 FLAIR. The UK Biobank team processed and quality-controlled the estimates of white matter volume, providing them as image-derived phenotypes to approved researchers. A comprehensive summary of the data acquisition protocols and preprocessing procedures is available at https://biobank.ctsu.ox.ac.uk/crystal/crystal/docs/brain_mri.pdf.

### Covariates

Based on prior studies on sunlight and cognitive function, the following factors were identified as potential confounding variables: age, sex(male or female), Townsend Deprivation Index (TDI), years of education(10-years, 13-years, 15-years, 19-years, or 20-years),[32] employment status(yes or no), physical activity(low, moderate, high), body mass index (BMI), smoking status(never, previous, or current), alcohol drinker status(never, previous, or current), skin color(very fair, fair, light olive, dark olive, brown, black), use of sun/UV protection(never/rarely, sometimes, most of the time, always, do not go out in sunshine), history of fractures in the past 5 years(yes or no), vitamin D supplementation(yes or no), sleep duration(7-8 hours or not), history of hypertension(yes or no), history of stroke(yes or no), history of coronary heart disease(yes or no), and history of diabetes(yes or no).

In addition, we adjusted for PM2.5 and PRS for Alzheimer’s disease (AD-PRS) to control environmental pollution factor and genetic factor. The assessment centers were adjusted to control the impact of the brain scanning device. The detailed definitions of hypertension, stroke, coronary heart disease, and diabetes can be found in Table S1.

## Statistical analyses

Normally distributed variables were presented as mean (standard deviation), non-normally distributed variables as median (interquartile range), and categorical variables as numbers (percentages).

Participants were stratified into three groups based on the tertiles of sunlight exposure time (Tertile 1, Tertile 2 Tertile 3), with the group having the lowest sunlight exposure time considered as the reference group. Linear regression analysis was employed to compare the differences in brain structural markers among different sunlight exposure time groups. In the stratified analysis, the subjects were divided into subgroups based on sex, age (< 60, >= 60), and disease history (hypertension, stroke, and diabetes). Within each subgroup, we analyzed the relationships between sunlight exposure time and brain structural markers. Additionally, we treated sunlight exposure time as a continuous variable and employed the “plotRCS” package for restricted cubic splines to examine the dose-response relationship between sunlight exposure time and brain structural markers. Given variations in daylight duration between seasons, we also separately examined the dose-response relationships for summer and winter.

In sensitivity analyses, we separately excluded participants who developed dementia in the first 5 years of follow-up and 10 years of follow-up, to control for potential reverse causality. Participants with a history of hypertension, diabetes, or stroke at baseline were excluded, and then repeating the primary analysis in a relatively healthy population. The relationships between sunlight exposure time and different cognitive domains were also analyzed. (Table S2)[33]

The statistical analyses were conducted in R version 4.2.3, and statistical significance was set at the *p*-value < 0.05 for two-tailed tests.

## Results

A total of 27,474 participants(mean age 55.01 ± 7.57years) who completed brain scan were included in baseline characterization analysis. (Table 1) Compared to the group with shorter sunlight exposure time, the group with longer time tends to be older, more likely to consist of males, engage in high level of physical activities, and have appropriate sleep duration.

**Table 1.**
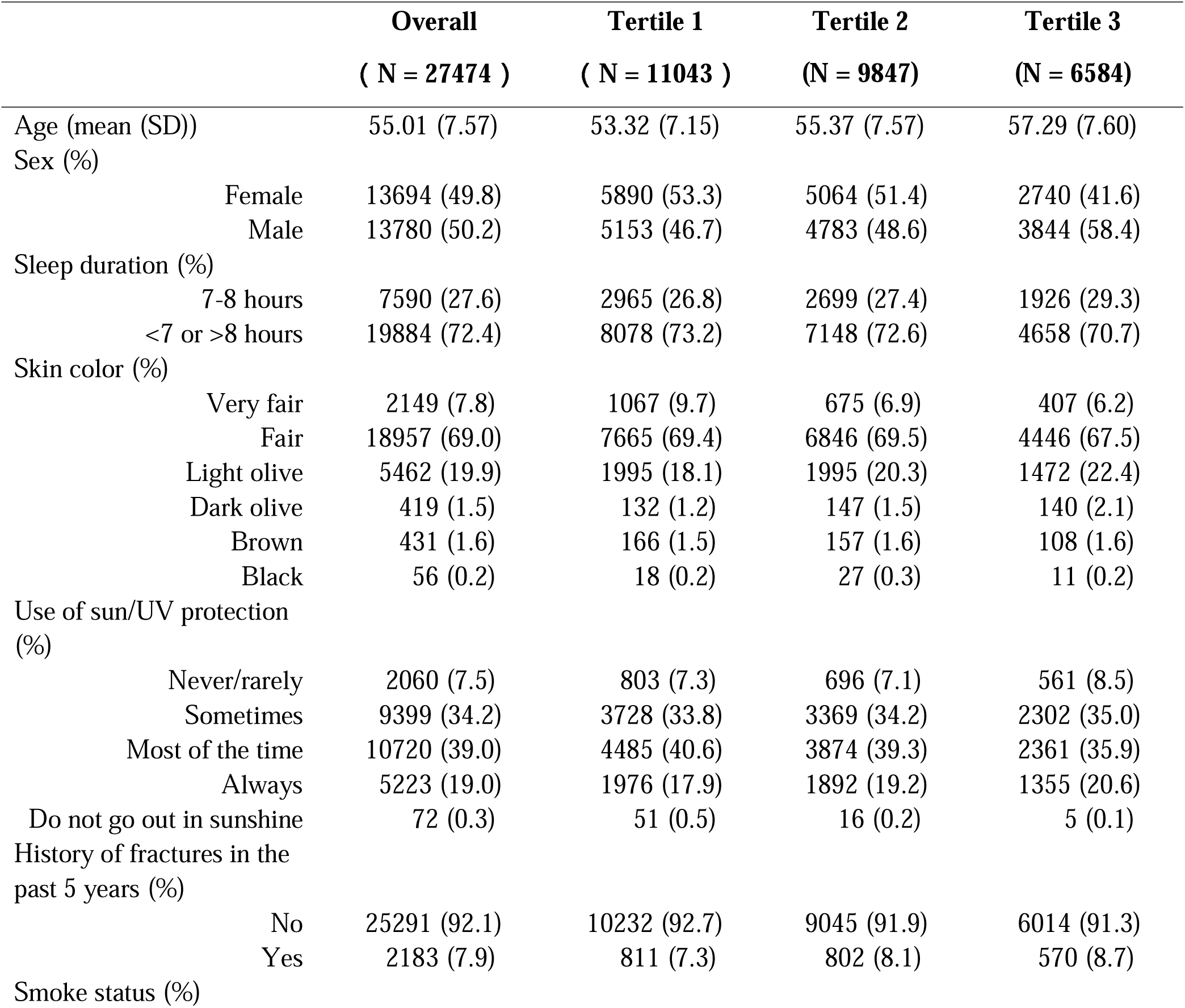
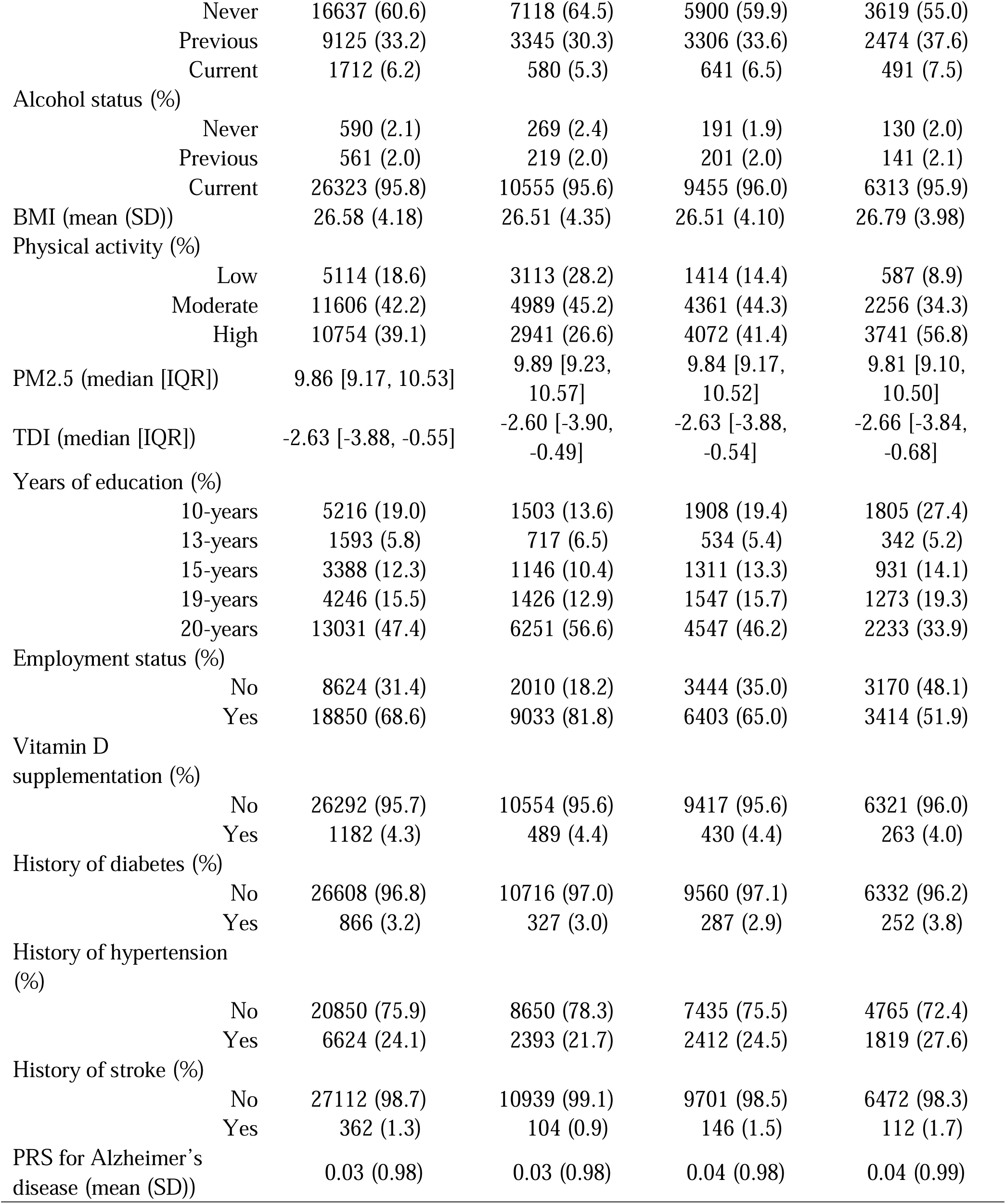
Participant Characteristics

|  | **Overall**<br>**( N = 27474 )** | **Tertile 1**<br>**( N = 11043 )** | **Tertile 2**<br>**(N = 9847)** | **Tertile 3**<br>**(N = 6584)** |
| --- | --- | --- | --- | --- |
| Age (mean (SD)) | 55.01 (7.57) | 53.32 (7.15) | 55.37 (7.57) | 57.29 (7.60) |
| Sex (%) |  |  |  |  |
| Female | 13694 (49.8) | 5890 (53.3) | 5064 (51.4) | 2740 (41.6) |
| Male | 13780 (50.2) | 5153 (46.7) | 4783 (48.6) | 3844 (58.4) |
| Sleep duration (%) |  |  |  |  |
| 7-8 hours | 7590 (27.6) | 2965 (26.8) | 2699 (27.4) | 1926 (29.3) |
| <7 or >8 hours | 19884 (72.4) | 8078 (73.2) | 7148 (72.6) | 4658 (70.7) |
| Skin color (%) |  |  |  |  |
| Very fair | 2149 (7.8) | 1067 (9.7) | 675 (6.9) | 407 (6.2) |
| Fair | 18957 (69.0) | 7665 (69.4) | 6846 (69.5) | 4446 (67.5) |
| Light olive | 5462 (19.9) | 1995 (18.1) | 1995 (20.3) | 1472 (22.4) |
| Dark olive | 419 (1.5) | 132 (1.2) | 147 (1.5) | 140 (2.1) |
| Brown | 431 (1.6) | 166 (1.5) | 157 (1.6) | 108 (1.6) |
| Black | 56 (0.2) | 18 (0.2) | 27 (0.3) | 11 (0.2) |
| Use of sun/UV protection (%) |  |  |  |  |
| Never/rarely | 2060 (7.5) | 803 (7.3) | 696 (7.1) | 561 (8.5) |
| Sometimes | 9399 (34.2) | 3728 (33.8) | 3369 (34.2) | 2302 (35.0) |
| Most of the time | 10720 (39.0) | 4485 (40.6) | 3874 (39.3) | 2361 (35.9) |
| Always | 5223 (19.0) | 1976 (17.9) | 1892 (19.2) | 1355 (20.6) |
| Do not go out in sunshine | 72 (0.3) | 51 (0.5) | 16 (0.2) | 5 (0.1) |
| History of fractures in the past 5 years (%) |  |  |  |  |
| No | 25291 (92.1) | 10232 (92.7) | 9045 (91.9) | 6014 (91.3) |
| Yes | 2183 (7.9) | 811 (7.3) | 802 (8.1) | 570 (8.7) |
| Smoke status (%) |  |  |  |  |

|  |  |  |  |  |  |
| --- | --- | --- | --- | --- | --- |
|  | Never | 16637 (60.6) | 7118 (64.5) | 5900 (59.9) | 3619 (55.0) |
|  | Previous | 9125 (33.2) | 3345 (30.3) | 3306 (33.6) | 2474 (37.6) |
|  | Current | 1712 (6.2) | 580 (5.3) | 641 (6.5) | 491 (7.5) |
| Alcohol status (%) |  |  |  |  |  |
|  | Never | 590 (2.1) | 269 (2.4) | 191 (1.9) | 130 (2.0) |
|  | Previous | 561 (2.0) | 219 (2.0) | 201 (2.0) | 141 (2.1) |
|  | Current | 26323 (95.8) | 10555 (95.6) | 9455 (96.0) | 6313 (95.9) |
| BMI (mean (SD)) |  | 26.58 (4.18) | 26.51 (4.35) | 26.51 (4.10) | 26.79 (3.98) |
| Physical activity (%) |  |  |  |  |  |
|  | Low | 5114 (18.6) | 3113 (28.2) | 1414 (14.4) | 587 (8.9) |
|  | Moderate | 11606 (42.2) | 4989 (45.2) | 4361 (44.3) | 2256 (34.3) |
|  | High | 10754 (39.1) | 2941 (26.6) | 4072 (41.4) | 3741 (56.8) |
| PM2.5 (median [IQR]) |  | 9.86 [9.17, 10.53] | 9.89 [9.23, 10.57] | 9.84 [9.17, 10.52] | 9.81 [9.10, 10.50] |
| TDI (median [IQR]) |  | -2.63 [-3.88, -0.55] | -2.60 [-3.90, -0.49] | -2.63 [-3.88, -0.54] | -2.66 [-3.84, -0.68] |
| Years of education (%) |  |  |  |  |  |
|  | 10-years | 5216 (19.0) | 1503 (13.6) | 1908 (19.4) | 1805 (27.4) |
|  | 13-years | 1593 (5.8) | 717 (6.5) | 534 (5.4) | 342 (5.2) |
|  | 15-years | 3388 (12.3) | 1146 (10.4) | 1311 (13.3) | 931 (14.1) |
|  | 19-years | 4246 (15.5) | 1426 (12.9) | 1547 (15.7) | 1273 (19.3) |
|  | 20-years | 13031 (47.4) | 6251 (56.6) | 4547 (46.2) | 2233 (33.9) |
| Employment status (%) |  |  |  |  |  |
|  | No | 8624 (31.4) | 2010 (18.2) | 3444 (35.0) | 3170 (48.1) |
|  | Yes | 18850 (68.6) | 9033 (81.8) | 6403 (65.0) | 3414 (51.9) |
| Vitamin D supplementation (%) |  |  |  |  |  |
|  | No | 26292 (95.7) | 10554 (95.6) | 9417 (95.6) | 6321 (96.0) |
|  | Yes | 1182 (4.3) | 489 (4.4) | 430 (4.4) | 263 (4.0) |
| History of diabetes (%) |  |  |  |  |  |
|  | No | 26608 (96.8) | 10716 (97.0) | 9560 (97.1) | 6332 (96.2) |
|  | Yes | 866 (3.2) | 327 (3.0) | 287 (2.9) | 252 (3.8) |
| History of hypertension (%) |  |  |  |  |  |
|  | No | 20850 (75.9) | 8650 (78.3) | 7435 (75.5) | 4765 (72.4) |
|  | Yes | 6624 (24.1) | 2393 (21.7) | 2412 (24.5) | 1819 (27.6) |
| History of stroke (%) |  |  |  |  |  |
|  | No | 27112 (98.7) | 10939 (99.1) | 9701 (98.5) | 6472 (98.3) |
|  | Yes | 362 (1.3) | 104 (0.9) | 146 (1.5) | 112 (1.7) |
| PRS for Alzheimer's disease (mean (SD)) |  | 0.03 (0.98) | 0.03 (0.98) | 0.04 (0.98) | 0.04 (0.99) |

## Main analysis

The association between sunlight exposure time and brain structural markers was presented in Table 2. Comparing to Tertile 1, prolong natural sunlight exposure time (Tertile 3) was associated with the volumes of total brain (β: -0.051, P < 0.001), white matter (β: -0.031, P = 0.023), gray matter (β: -0.067, P < 0.001), and white matter hyperintensities (β: 0.059, P < 0.001). Longer sunlight exposure time was associated with smaller subcortical volumes of thalamus (β: -0.060, P < 0.001), caudate (β: -0.040, P = 0.012), putamen (β: -0.031, P = 0.032), hippocampus (β: -0.046, P = 0.003), and accumbens (β: -0.041, P = 0.006). Similarly, prolonged sunlight exposure was associated with reduced gray matter volumes in the putamen (β: -0.060, P < 0.001), hippocampus (β: -0.043, P = 0.004), and amygdala (β: -0.073, P < 0.001).

**Table 2.** Table Association between sunlight exposure time and brain structural markers

|  | Tertile 2 |  |  | Tertile 3 |  |  |
| --- | --- | --- | --- | --- | --- | --- |
| | $\beta$ | SE | P | $\beta$ | SE | P |
| **Global measures** |  |  |  |  |  |  |
| Total brain volume | -0.025 | 0.011 | 0.026 | -0.051 | 0.013 | <0.001 |
| Volume of white matter | -0.014 | 0.012 | 0.222 | -0.031 | 0.014 | 0.023 |
| Volume of gray matter | -0.035 | 0.011 | 0.002 | -0.067 | 0.014 | <0.001 |
| Volume of white matter hyperintensities | 0.013 | 0.013 | 0.314 | 0.059 | 0.016 | <0.001 |
| **Subcortical regions** |  |  |  |  |  |  |
| Volume of thalamus | -0.029 | 0.012 | 0.014 | -0.060 | 0.014 | <0.001 |
| Volume of caudate | -0.030 | 0.013 | 0.025 | -0.040 | 0.016 | 0.012 |
| Volume of putamen | -0.022 | 0.012 | 0.065 | -0.031 | 0.014 | 0.032 |
| Volume of pallidum | -0.027 | 0.013 | 0.044 | -0.026 | 0.016 | 0.106 |
| Volume of hippocampus | -0.010 | 0.013 | 0.450 | -0.046 | 0.016 | 0.003 |
| Volume of amygdala | -0.003 | 0.014 | 0.819 | -0.031 | 0.016 | 0.054 |
| Volume of accumbens | -0.004 | 0.013 | 0.748 | -0.041 | 0.015 | 0.006 |
| **Regional gray matter volumes** |  |  |  |  |  |  |
| Volume of thalamus | -0.007 | 0.014 | 0.613 | -0.022 | 0.016 | 0.179 |
| Volume of caudate | -0.010 | 0.014 | 0.461 | 0.009 | 0.016 | 0.566 |
| Volume of putamen | -0.052 | 0.014 | 0.000 | -0.060 | 0.017 | <0.001 |
| Volume of pallidum | 0.001 | 0.014 | 0.960 | 0.018 | 0.017 | 0.276 |
| Volume of hippocampus | -0.018 | 0.013 | 0.154 | -0.043 | 0.015 | 0.004 |
| Volume of amygdala | -0.038 | 0.012 | 0.002 | -0.070 | 0.015 | <0.001 |

## Stratified analysis

The male brain structure appears to be more susceptible to the effects of sunlight exposure compared to females. (Table S3) Among males, we found that prolong sunlight exposure was negatively associated with total brain volume, gray matter volume, subcortical volumes of the thalamus and caudate, gray matter volumes of the putamen, hippocampus, and amygdala. It is also associated with an increase in the volume of white matter hyperintensity. In females, it was only associated with total brain volume, gray matter volume, subcortical volumes of the thalamus and hippocampus.

Compared to the group aged 60 and above, the group under 60 years old showed a broader range of correlations between sunlight exposure and brain structural markers. (Table S4) With longer sunlight exposure time, participants under 60 exhibited shrinkage in volumes of total brain, white matter, gray matter, and increase in volume of white matter hyperintensities. However, only a correlation with gray matter volume was found in the population aged 60 and above.

In the group of hypertension, prolong sunlight exposure time was associated with total brain volume, gray matter volume, white matter volume, subcortical volumes in thalamus and hippocampus, and the gray matter volumes in putamen, hippocampus and amygdala. However, no significant associations were observed in the stroke and diabetes individuals. (Table S5)

## Restricted cubic spline

The restricted cubic spline illustrates a dose-response relationship between sunlight exposure duration and the volume of brain structural markers. With daily sunlight exposure approximately exceeding 2 hours, we observed that total brain volume, gray matter volume, and volumes of certain subcortical regions decreased with prolonged sunlight exposure duration. (Figure 2 and Figure S1) When stratified by season, as sunlight exposure duration increases, the total brain volume, white matter volume, and gray matter volume decreased more pronounced in the summer compared to winter. (Figure 3 and Figure S2) Regardless of the season, sunlight exposure time is associated with an increase in white matter hyperintensity volume.

**Figure 2.**
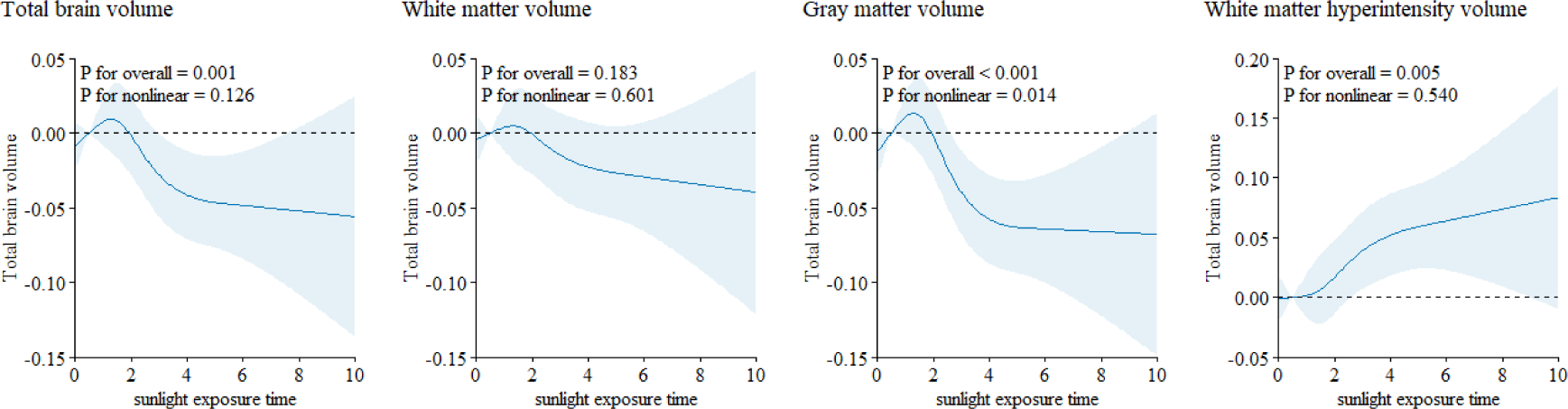
The restricted cubic spline of natural sunlight exposure with brain structure markers

**Figure 2.**
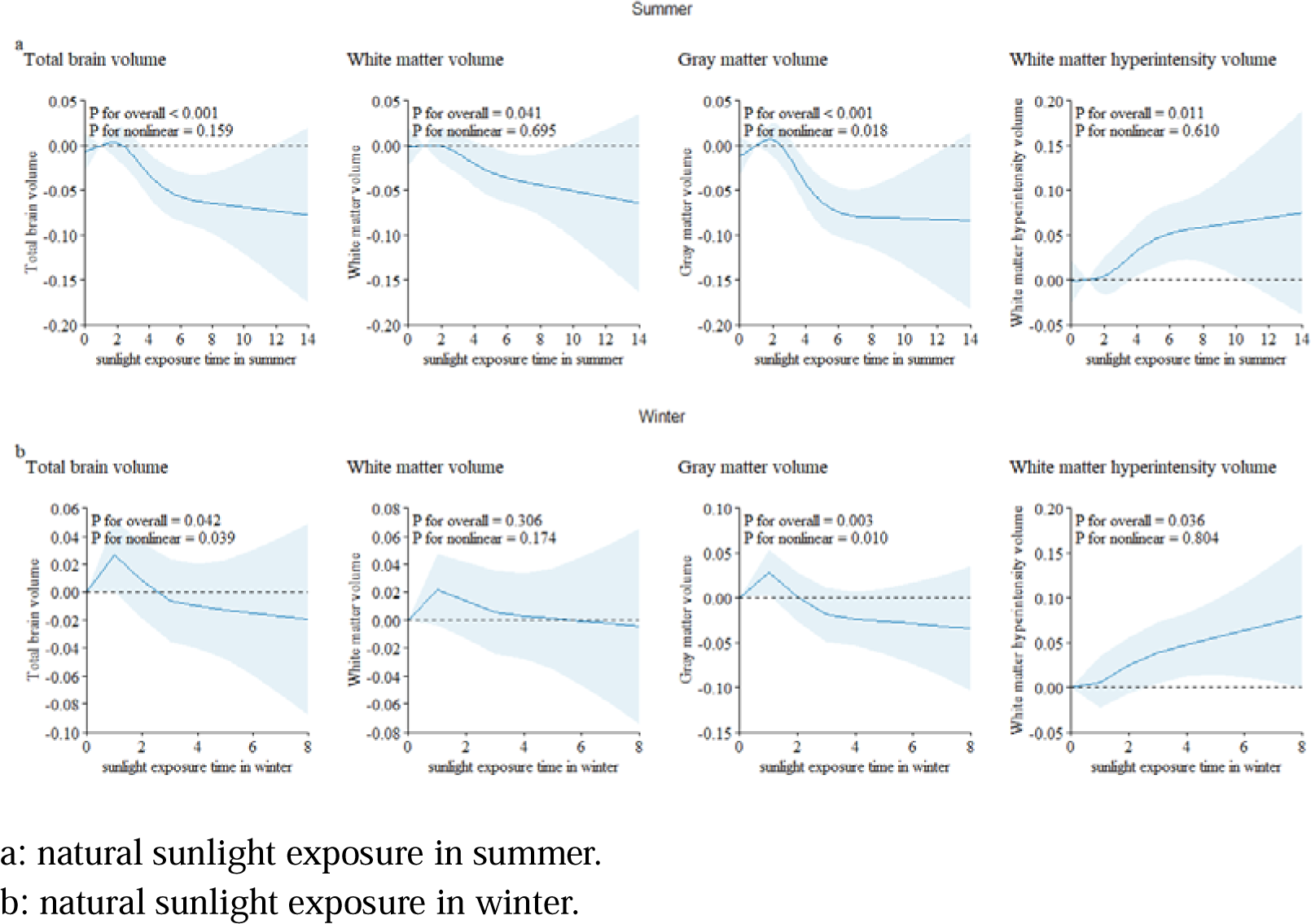
The restricted cubic spline of natural sunlight exposure with brain structure markers stratified by season.

## Sensitivity analysis

The sensitivity analyses results showed that our findings were robust. After excluding participants who developed dementia within the first 5 years and the first 10 years of follow-up, the results still consistent with the main results. (Table S6 and Table S7) Similar results were found between prolonged natural sunlight exposure and brain structural markers when baseline hypertension, diabetes, or stroke individuals were further removed. (Table S8) Prolong sunlight exposure time was associated with cognitive function tests. (Table S9) In terms of cognitive function, as the duration of sunlight exposure increased, performance in visual declarative memory, working memory, verbal and numerical reasoning, processing speed, executive function, vocabulary, and non-verbal reasoning declined.

## Discussion

We observed that prolonged exposure to natural sunlight may be associated with adverse brain structure. This association varies with age, gender, and season, with stronger negative correlations found in males, those under 60 years old, and during the summer. Additionally, prolonged exposure to sunlight is correlated with cognitive decline.

The mechanisms of sunlight-induced damage to brain structure are not fully understood and maybe the following two mechanisms: (1) Sunlight exposure causes an increase in core temperature, and then more blood flowing away from the brain, resulting in reduced cerebral blood flow, which in turn can cause damage to brain structure.[15–17] (2) The UV radiation in natural sunlight can damage immune cells in the body, triggering inflammatory responses that can lead to damage.[7, 9]

The relationship between natural sunlight exposure and change in brain structure, appear to be more extensive in the summer season, in individuals younger than 60 years old, and males. This can be attributed to higher temperature and stronger UV radiation during the summer in the United Kingdom.[34] Additionally, during the summer, people tend to expose more skin due to warmer weather and clothing choices, leading to increased UV exposure. Younger individuals tend to engage in outdoor activities, and research has found that among people above 20 years, the frequency of sunburn decreases with age.[35, 36] There are known structural and biological differences in the skin between sex.[37, 38] Compared to females, males tend to be more sensitive to UV radiation and may experience immune-suppression reactions more frequently.[39, 40] Conversely, the presence of estrogen in the female body may exert inhibitory effects on immune-suppression reactions.[41] Furthermore, males are generally less likely to use sun protection measures, resulting in greater sunlight exposure.[36]

The associations between sunlight exposure and brain structural markers are consistent with prospective studies in dementia populations. Ma, L.-Z., et al. found a “J-shaped” relationship between sunlight exposure and the development of dementia, and we observed that high-dose sunlight exposure may have a damaging effect on brain structural markers. [2] The finding regarding the association of natural sunlight exposure with cognition align with previous comparative studies conducted on worker populations. Exposure to sunlight has been observed to decrease attention allocation and vigilance. Under both temperate and tropical climate conditions, sunlight exposure has been shown to result in cognitive impairment.[42] Dementia is a slowly progressive condition, and the cognition changes we focused on occur earlier than the diagnosis of dementia.[43–46] Besides, research indicates that the atrophy of white matter may lead to cognitive impairment such as vascular dementia and other related conditions.[12, 47–49] Additionally, the atrophy of gray matter volume is also associated with the decline in cognition, such as in Alzheimer’s disease.[48, 50] Based on the association between natural sunlight and changes in brain structure, we hypothesize that brain structure may mediate the association between natural sunlight and cognition. However, further research is needed to confirm this hypothesis.

This study represents the first exploration of the associations and dose-response relationship between natural sunlight exposure and brain structure in the general population. We extensively adjusted for various potential confounding factors to control for influences from the environment, genetics, and other aspects. Furthermore, we conducted multi-level analyses stratified by season, age, sex, and four diseases to investigate variations among different subgroups. However, there are still some limitations. First, sunlight exposure time relied on self-reports from participants, which may introduce recall bias and subjective assessment. Second, the observational nature of this study prevents us from establishing causality. Third, the associations between sunlight exposure and brain structure were not observed in groups with specific diseases due to the relatively small number of participants with those conditions. Fourth, the participants in this study were primarily white individuals from high-latitude regions, which may limit the generalizability of the findings to other regions and ethnicities.

## Conclusions

In conclusion, this study reveals an association between prolonged exposure to natural sunlight and adverse changes in brain structure, providing novel insights into the potential impact of light exposure on human health. The findings highlight the need for further in-depth investigations to elucidate the specific mechanisms and physiological foundations underlying this relationship. Understanding the intricacies of how natural sunlight affects brain structure is crucial for advancing our knowledge of the broader implications for human well-being.

## Data Availability

Because this study used UK Biobank data, it is not possible to provide all the data.

## Author Contributions

Prof. Dongfeng Zhang had full access to all of the data in the study and takes responsibility for the integrity of the data and the accuracy of the data analysis.

## Concept and design

Huihui Li, Fusheng Cui.

## Acquisition, analysis, or interpretation of data

All authors.

## Drafting of the manuscript

Huihui Li, Fusheng Cui.

## Critical revision of the manuscript for important intellectual content

Dongfeng Zhang, Weijing Wang.

## Statistical analysis

Fusheng Cui, Tong Wang, Weijing Wang.

## Obtained funding

Dongfeng Zhang, Weijing Wang.

## Administrative, technical, or material support

Dongfeng Zhang, Weijing Wang, Tong Wang.

## Supervision

Dongfeng Zhang, Weijing Wang, Tong Wang.

## Conflict of Interest Disclosures

None reported.

## Funding/Support

This study was supported by the National Natural Science Foundation of China (82073641).

## Role of the Funder/Sponsor

The funding organizations had no role in the design and conduct of the study; collection, management, analysis, and interpretation of the data; preparation, review, or approval of the manuscript; and decision to submit the manuscript for publication.

## Author contributions

D.Z. and H.L. designed research; H.L. performed research; H.L, F.C. analyzed data; H.L., F.C, T.W., W.W., and D.Z. wrote the first draft of the paper; H.L., F.C, T.W., W.W., and D.Z. edited the paper; H.L., F.C, T.W., W.W., and D.Z. wrote the paper.

## Funding/Support

This study was supported by the National Natural Science Foundation of China (82073641).

## Competing interests

The authors declare that they have no competing interests.

## Preprints

This manuscript has been submitted as preprint at MedRxiv: 10.1101/2023.10.12.23296944.

## Acknowledgements

This study utilized data from the UK Biobank and was approved by the UK Biobank (proposal 95715). The authors gratefully thank all the participants and professionals contributing to the UK Biobank.

## Abbreviations

UV: ultraviolet
TDI: Townsend Deprivation Index
BMI: body mass index
AD-PRS: PRS for Alzheimer’s disease

## Notes

### Competing Interest Statement

The authors have declared no competing interest.

### Funding Statement

This study was funded by the National Natural Science Foundation of China (82073641).

### Summary of Updates

Sunlight exposure time was grouped by triadile, and dose-response relationship analysis was added

