## Supplement for "Associations between natural sunlight exposure and brain structural markers: a prospective study in the UK Biobank"

### Table S1. Definitions of hypertension, stroke, coronary heart disease, and diabetes

| **Diseases** | **Self-reported Information** | **ICD-9 information** | **ICD-10 information** | **OPCS-4 information** |
| --- | --- | --- | --- | --- |
| Hypertension | 2966, 6150 (4), 6153 (2), 6177 (2), 20002 (1065, 1072), 20008, 20009, 20010, 20011 | 41271 (401-405) 41281 | 41270 (I10-I13, I15, O10), 41280, 131292, 131290, 131288, 131286, 131294, 132180, 131295, 131293, 131291, 131289, 131287, 132181 |  |
| Stroke | 6150 (3), 4056, 20002 (1081, 1491, 1583, 1086), 20008, 20009, 20010, 20011 | 41271 (3361, 3623, 430, 431, 4329, 4330, 4331, 4332,4333, 4338, 4339, 434, 436), 41281 | 41270 (I60, I61, I629, I63, I64, I678, I690, I693, G951, H341, H342, S066), 41280, 131378, 131376, 131374, 131372, 131370, 131368, 131366, 131364, 131362, 131360, 131180, 131379, 131377, 131375, 131373, 131371, 131369, 131367, 131365, 131363, 131361, 131181 | 41272 (A052- A054, L351, L353, L343), 41282 |
| Diabetes | 2443 (1), 2976, 6153 (3), 6177(3), 20002 (1220, 1222, 1223), 20008, 20009 | 41271 (250, 3572, 3620), 41281 | 41270 (E10-E14, G590, G632, H280, H360, M142, N083) 41280, 130714, 130712, 130710, 130708, 130706, 130715, 130713, 130711, 130709, 130707 |  |

ICD: International Classification of Diseases

the Office of Population Censuses and Surveys Classification of Interventions and Procedures, version 4

### Table S2. Cognitive domain, definition, and Field ID details for cognitive function tests

| **Cognitive function tests** | **Cognitive domain** | **definition** | **Field ID** |
| --- | --- | --- | --- |
| Prospective memory test | prospective memory | the initial chose was correct | 20118 |
| Pairs matching test | visual declarative memory | number of incorrect matches in round | 399 |
| Numeric memory test | working memory | maximum digits remembered correctly | 4282 |
| Fluid Intelligence test | verbal and numerical reasoning | fluid intelligence score | 20016 |
| Reaction time test | processing speed | mean time to correctly identify matches | 20023 |
| Trail making test 1 | executive function | duration to complete numeric path | 6348 |
| Trail making test 2 | executive function | duration to complete alphanumeric path | 6350 |
| Symbol digit substitution test | processing speed | number of symbol digit matches made correctly | 23324 |
| Picture vocabulary test | vocabulary | vocabulary level | 6364 |
| Paired associate learning test | visual declarative memory | number of word pairs correctly associated | 20197 |
| Matrix pattern completion test | non-verbal reasoning | number of puzzles correctly solved | 6373 |
| Tower rearranging test | executive function | number of puzzles correct | 21004 |

### Table S3. Table Association between sunlight exposure time and brain structural markers stratified by sex (compare to Tertile 1).

|  |  | **Tertile 2** |  |  | **Tertile 3** |  |
| --- | --- | --- | --- | --- | --- | --- |
|  | ***β*** | **SE** | **P** | ***β*** | **SE** | **P** |
| **Global measures** |  |  |  |  |  |  |
| Total brain volume |  |  |  |  |  |  |
| Male | -0.037 | 0.019 | 0.056 | -0.064 | 0.022 | 0.004 |
| Female | -0.022 | 0.018 | 0.231 | -0.058 | 0.023 | 0.011 |
| Volume of white matter |  |  |  |  |  |  |
| Male | -0.021 | 0.020 | 0.299 | -0.040 | 0.023 | 0.082 |
| Female | -0.013 | 0.019 | 0.502 | -0.041 | 0.024 | 0.082 |
| Volume of gray matter |  |  |  |  |  |  |
| Male | -0.048 | 0.018 | 0.009 | -0.079 | 0.021 | <0.001 |
| Female | -0.028 | 0.017 | 0.106 | -0.067 | 0.022 | 0.002 |
| Volume of white matter hyperintensities |  |  |  |  |  |  |
| Male | 0.012 | 0.019 | 0.536 | 0.065 | 0.022 | 0.003 |
| Female | 0.016 | 0.018 | 0.376 | 0.043 | 0.023 | 0.056 |
| **Subcortical regions** |  |  |  |  |  |  |
| Volume of thalamus |  |  |  |  |  |  |
| Male | -0.032 | 0.018 | 0.083 | -0.057 | 0.021 | 0.006 |
| Female | -0.033 | 0.018 | 0.072 | -0.065 | 0.023 | 0.004 |
| Volume of caudate |  |  |  |  |  |  |
| Male | -0.062 | 0.020 | 0.002 | -0.059 | 0.023 | 0.011 |
| Female | 0.001 | 0.020 | 0.950 | -0.022 | 0.024 | 0.357 |
| Volume of putamen |  |  |  |  |  |  |
| Male | -0.031 | 0.019 | 0.103 | -0.031 | 0.022 | 0.156 |
| Female | -0.018 | 0.019 | 0.336 | -0.033 | 0.023 | 0.154 |
| Volume of pallidum |  |  |  |  |  |  |
| Male | -0.048 | 0.020 | 0.016 | -0.031 | 0.023 | 0.178 |
| Female | -0.009 | 0.020 | 0.652 | -0.012 | 0.024 | 0.616 |
| Volume of hippocampus |  |  |  |  |  |  |
| Male | -0.021 | 0.019 | 0.282 | -0.030 | 0.022 | 0.170 |
| Female | 0.001 | 0.019 | 0.975 | -0.055 | 0.024 | 0.020 |
| Volume of amygdala |  |  |  |  |  |  |
| Male | <0.001 | 0.021 | 0.982 | -0.038 | 0.024 | 0.109 |
| Female | -0.008 | 0.020 | 0.668 | -0.019 | 0.025 | 0.432 |
| Volume of accumbens |  |  |  |  |  |  |
| Male | -0.022 | 0.018 | 0.230 | -0.034 | 0.021 | 0.097 |
| Female | 0.014 | 0.018 | 0.422 | -0.040 | 0.022 | 0.076 |
| **Regional gray matter volumes** |  |  |  |  |  |  |
| Volume of thalamus |  |  |  |  |  |  |
| Male | 0.013 | 0.021 | 0.531 | -0.030 | 0.024 | 0.196 |
| Female | -0.027 | 0.020 | 0.169 | -0.018 | 0.025 | 0.458 |
| Volume of caudate |  |  |  |  |  |  |
| Male | -0.024 | 0.020 | 0.238 | -0.006 | 0.023 | 0.797 |
| Female | 0.006 | 0.019 | 0.753 | 0.026 | 0.024 | 0.284 |
| Volume of putamen |  |  |  |  |  |  |
| Male | -0.068 | 0.020 | 0.001 | -0.074 | 0.023 | 0.002 |
| Female | -0.038 | 0.020 | 0.057 | -0.032 | 0.024 | 0.187 |
| Volume of pallidum |  |  |  |  |  |  |
| Male | 0.015 | 0.020 | 0.466 | 0.029 | 0.023 | 0.209 |
| Female | -0.013 | 0.019 | 0.511 | 0.001 | 0.024 | 0.968 |
| Volume of hippocampus |  |  |  |  |  |  |
| Male | -0.035 | 0.019 | 0.073 | -0.050 | 0.022 | 0.025 |
| Female | -0.004 | 0.019 | 0.839 | -0.037 | 0.024 | 0.113 |
| Volume of amygdala |  |  |  |  |  |  |
| Male | -0.052 | 0.019 | 0.005 | -0.090 | 0.021 | <0.001 |
| Female | -0.029 | 0.018 | 0.111 | -0.043 | 0.022 | 0.057 |

### Table S4. Table Association between sunlight exposure time and brain structural markers stratified by age (compare to Tertile 1).

|  |  | **Tertile 2** |  |  | **Tertile 3** |  |
| --- | --- | --- | --- | --- | --- | --- |
|  | ***β*** | **SE** | **P** | ***β*** | **SE** | **P** |
| **Global measures** |  |  |  |  |  |  |
| Total brain volume | -0.032 | 0.014 | 0.018 | -0.058 | 0.017 | 0.001 |
| <60 | -0.002 | 0.022 | 0.919 | -0.028 | 0.024 | 0.237 |
| >=60 |  |  |  |  |  |  |
| Volume of white matter |  |  |  |  |  |  |
| <60 | -0.024 | 0.014 | 0.080 | -0.044 | 0.017 | 0.011 |
| >=60 | 0.019 | 0.022 | 0.390 | 0.001 | 0.024 | 0.972 |
| Volume of gray matter |  |  |  |  |  |  |
| <60 | -0.039 | 0.014 | 0.006 | -0.069 | 0.018 | <0.001 |
| >=60 | -0.026 | 0.023 | 0.248 | -0.058 | 0.025 | 0.020 |
| Volume of white matter hyperintensities |  |  |  |  |  |  |
| <60 | 0.011 | 0.016 | 0.483 | 0.062 | 0.020 | 0.002 |
| >=60 | 0.020 | 0.026 | 0.437 | 0.053 | 0.028 | 0.063 |
| **Subcortical regions** |  |  |  |  |  |  |
| Volume of thalamus |  |  |  |  |  |  |
| <60 | -0.045 | 0.015 | 0.002 | -0.065 | 0.018 | <0.001 |
| >=60 | 0.015 | 0.024 | 0.532 | -0.028 | 0.026 | 0.282 |
| Volume of caudate |  |  |  |  |  |  |
| <60 | -0.035 | 0.016 | 0.027 | -0.041 | 0.020 | 0.037 |
| >=60 | -0.022 | 0.026 | 0.387 | -0.032 | 0.028 | 0.243 |
| Volume of putamen |  |  |  |  |  |  |
| <60 | -0.034 | 0.015 | 0.020 | -0.052 | 0.018 | 0.004 |
| >=60 | 0.010 | 0.024 | 0.681 | 0.026 | 0.026 | 0.327 |
| Volume of pallidum |  |  |  |  |  |  |
| <60 | -0.047 | 0.016 | 0.003 | -0.044 | 0.020 | 0.026 |
| >=60 | 0.024 | 0.026 | 0.358 | 0.037 | 0.028 | 0.182 |
| Volume of hippocampus |  |  |  |  |  |  |
| <60 | -0.034 | 0.016 | 0.034 | -0.046 | 0.020 | 0.023 |
| >=60 | 0.052 | 0.026 | 0.045 | <0.001 | 0.028 | 0.986 |
| Volume of amygdala |  |  |  |  |  |  |
| <60 | -0.013 | 0.016 | 0.431 | -0.012 | 0.020 | 0.534 |
| >=60 | 0.013 | 0.026 | 0.601 | -0.052 | 0.028 | 0.063 |
| Volume of accumbens |  |  |  |  |  |  |
| <60 | -0.014 | 0.016 | 0.391 | -0.057 | 0.020 | 0.004 |
| >=60 | 0.033 | 0.026 | 0.202 | 0.014 | 0.028 | 0.620 |
| **Regional gray matter volumes** |  |  |  |  |  |  |
| Volume of thalamus |  |  |  |  |  |  |
| <60 | -0.021 | 0.016 | 0.188 | -0.029 | 0.020 | 0.156 |
| >=60 | 0.036 | 0.025 | 0.159 | 0.010 | 0.027 | 0.724 |
| Volume of caudate |  |  |  |  |  |  |
| <60 | -0.024 | 0.016 | 0.138 | -0.003 | 0.021 | 0.870 |
| >=60 | 0.007 | 0.027 | 0.785 | 0.017 | 0.029 | 0.558 |
| Volume of putamen |  |  |  |  |  |  |
| <60 | -0.063 | 0.016 | <0.001 | -0.055 | 0.021 | 0.007 |
| >=60 | -0.037 | 0.027 | 0.166 | -0.042 | 0.029 | 0.139 |
| Volume of pallidum |  |  |  |  |  |  |
| <60 | -0.007 | 0.017 | 0.685 | 0.017 | 0.021 | 0.416 |
| >=60 | 0.010 | 0.027 | 0.696 | 0.015 | 0.029 | 0.600 |
| Volume of hippocampus |  |  |  |  |  |  |
| <60 | -0.042 | 0.015 | 0.006 | -0.043 | 0.019 | 0.026 |
| >=60 | 0.045 | 0.025 | 0.072 | -0.001 | 0.027 | 0.974 |
| Volume of amygdala |  |  |  |  |  |  |
| <60 | -0.045 | 0.015 | 0.003 | -0.065 | 0.019 | 0.001 |
| >=60 | -0.024 | 0.025 | 0.329 | -0.059 | 0.027 | 0.027 |

### Table S5. Table Association between sunlight exposure time and brain structural markers stratified by diseases (compare to Tertile 1).

|  |  | **Tertile 2** |  |  | **Tertile 3** |  |
| --- | --- | --- | --- | --- | --- | --- |
|  | ***β*** | **SE** | **P** | ***β*** | **SE** | **P** |
| **Global measures** |  |  |  |  |  |  |
| Total brain volume |  |  |  |  |  |  |
| Hypertension | -0.053 | 0.024 | 0.028 | -0.081 | 0.028 | 0.003 |
| Diabetes | 0.016 | 0.068 | 0.808 | 0.017 | 0.075 | 0.817 |
| Stroke | 0.040 | 0.106 | 0.708 | 0.152 | 0.116 | 0.190 |
| Volume of white matter |  |  |  |  |  |  |
| Hypertension | -0.030 | 0.024 | 0.223 | -0.056 | 0.028 | 0.045 |
| Diabetes | 0.050 | 0.069 | 0.465 | 0.097 | 0.077 | 0.208 |
| Stroke | 0.012 | 0.108 | 0.910 | 0.181 | 0.119 | 0.128 |
| Volume of gray matter |  |  |  |  |  |  |
| Hypertension | -0.072 | 0.024 | 0.003 | -0.099 | 0.028 | <0.001 |
| Diabetes | -0.023 | 0.068 | 0.732 | -0.072 | 0.075 | 0.338 |
| Stroke | 0.063 | 0.107 | 0.559 | 0.099 | 0.118 | 0.403 |
| Volume of white matter hyperintensities |  |  |  |  |  |  |
| Hypertension | -0.009 | 0.028 | 0.753 | 0.029 | 0.032 | 0.365 |
| Diabetes | -0.046 | 0.077 | 0.547 | 0.018 | 0.086 | 0.830 |
| Stroke | -0.129 | 0.128 | 0.314 | -0.085 | 0.140 | 0.545 |
| **Subcortical regions** |  |  |  |  |  |  |
| Volume of thalamus |  |  |  |  |  |  |
| Hypertension | -0.047 | 0.025 | 0.066 | -0.099 | 0.029 | 0.001 |
| Diabetes | 0.151 | 0.071 | 0.033 | 0.084 | 0.079 | 0.288 |
| Stroke | 0.122 | 0.119 | 0.308 | 0.095 | 0.131 | 0.465 |
| Volume of caudate |  |  |  |  |  |  |
| Hypertension | -0.016 | 0.028 | 0.575 | -0.024 | 0.032 | 0.456 |
| Diabetes | 0.001 | 0.078 | 0.990 | 0.027 | 0.087 | 0.758 |
| Stroke | 0.129 | 0.125 | 0.303 | 0.097 | 0.137 | 0.479 |
| Volume of putamen |  |  |  |  |  |  |
| Hypertension | -0.004 | 0.026 | 0.884 | -0.016 | 0.029 | 0.581 |
| Diabetes | -0.024 | 0.071 | 0.734 | -0.049 | 0.080 | 0.540 |
| Stroke | 0.066 | 0.121 | 0.585 | 0.177 | 0.132 | 0.183 |
| Volume of pallidum |  |  |  |  |  |  |
| Hypertension | 0.005 | 0.028 | 0.864 | -0.012 | 0.032 | 0.713 |
| Diabetes | 0.003 | 0.077 | 0.969 | 0.041 | 0.086 | 0.635 |
| Stroke | 0.099 | 0.126 | 0.434 | 0.034 | 0.138 | 0.804 |
| Volume of hippocampus |  |  |  |  |  |  |
| Hypertension | 0.001 | 0.027 | 0.971 | -0.071 | 0.031 | 0.023 |
| Diabetes | 0.046 | 0.076 | 0.546 | 0.035 | 0.085 | 0.678 |
| Stroke | 0.013 | 0.126 | 0.916 | -0.064 | 0.138 | 0.642 |
| Volume of amygdala |  |  |  |  |  |  |
| Hypertension | -0.012 | 0.028 | 0.660 | -0.050 | 0.032 | 0.127 |
| Diabetes | -0.009 | 0.080 | 0.907 | 0.037 | 0.090 | 0.682 |
| Stroke | 0.075 | 0.128 | 0.558 | 0.070 | 0.140 | 0.618 |
| Volume of accumbens |  |  |  |  |  |  |
| Hypertension | 0.002 | 0.027 | 0.955 | -0.030 | 0.031 | 0.332 |
| Diabetes | 0.038 | 0.074 | 0.606 | 0.052 | 0.082 | 0.525 |
| Stroke | 0.157 | 0.121 | 0.193 | 0.093 | 0.132 | 0.482 |
| **Regional gray matter volumes** |  |  |  |  |  |  |
| Volume of thalamus |  |  |  |  |  |  |
| Hypertension | -0.006 | 0.028 | 0.823 | -0.038 | 0.032 | 0.240 |
| Diabetes | 0.075 | 0.078 | 0.336 | 0.120 | 0.087 | 0.169 |
| Stroke | 0.021 | 0.124 | 0.866 | 0.006 | 0.136 | 0.967 |
| Volume of caudate |  |  |  |  |  |  |
| Hypertension | -0.025 | 0.029 | 0.378 | -0.005 | 0.033 | 0.868 |
| Diabetes | 0.002 | 0.080 | 0.983 | 0.022 | 0.089 | 0.803 |
| Stroke | -0.059 | 0.127 | 0.640 | 0.140 | 0.139 | 0.315 |
| Volume of putamen |  |  |  |  |  |  |
| Hypertension | -0.027 | 0.029 | 0.343 | -0.080 | 0.033 | 0.016 |
| Diabetes | -0.078 | 0.080 | 0.333 | -0.079 | 0.089 | 0.378 |
| Stroke | 0.012 | 0.132 | 0.926 | -0.069 | 0.145 | 0.635 |
| Volume of pallidum |  |  |  |  |  |  |
| Hypertension | 0.030 | 0.029 | 0.307 | 0.029 | 0.033 | 0.380 |
| Diabetes | -0.011 | 0.082 | 0.895 | -0.023 | 0.091 | 0.799 |
| Stroke | 0.119 | 0.126 | 0.347 | 0.050 | 0.138 | 0.717 |
| Volume of hippocampus |  |  |  |  |  |  |
| Hypertension | -0.014 | 0.027 | 0.597 | -0.084 | 0.031 | 0.006 |
| Diabetes | 0.085 | 0.075 | 0.252 | -0.016 | 0.083 | 0.844 |
| Stroke | 0.010 | 0.123 | 0.932 | -0.012 | 0.135 | 0.928 |
| Volume of amygdala |  |  |  |  |  |  |
| Hypertension | -0.057 | 0.026 | 0.029 | -0.112 | 0.030 | <0.001 |
| Diabetes | -0.049 | 0.071 | 0.491 | -0.073 | 0.079 | 0.356 |
| Stroke | 0.068 | 0.119 | 0.570 | 0.132 | 0.131 | 0.315 |

### Table S6. Association between sunlight exposure time and brain structural markers after exclude dementia incidents at first five years of follow-up. (N = 27,471) (compare to Tertile 1)

|  |  | **Tertile 2** |  |  | **Tertile 3** |  |
| --- | --- | --- | --- | --- | --- | --- |
|  | ***β*** | **SE** | **P** | ***β*** | **SE** | **P** |
| **Global measures** |  |  |  |  |  |  |
| Total brain volume | -0.025 | 0.011 | 0.026 | -0.051 | 0.013 | <0.001 |
| Volume of white matter | -0.014 | 0.012 | 0.219 | -0.031 | 0.014 | 0.022 |
| Volume of gray matter | -0.035 | 0.011 | 0.002 | -0.067 | 0.014 | <0.001 |
| Volume of white matter hyperintensities | 0.013 | 0.013 | 0.317 | 0.059 | 0.016 | <0.001 |
| **Subcortical regions** |  |  |  |  |  |  |
| Volume of thalamus | -0.029 | 0.012 | 0.014 | -0.060 | 0.014 | <0.001 |
| Volume of caudate | -0.030 | 0.013 | 0.025 | -0.040 | 0.016 | 0.012 |
| Volume of putamen | -0.022 | 0.012 | 0.066 | -0.031 | 0.014 | 0.033 |
| Volume of pallidum | -0.027 | 0.013 | 0.044 | -0.026 | 0.016 | 0.106 |
| Volume of hippocampus | -0.010 | 0.013 | 0.451 | -0.046 | 0.016 | 0.003 |
| Volume of amygdala | -0.003 | 0.014 | 0.814 | -0.031 | 0.016 | 0.054 |
| Volume of accumbens | -0.004 | 0.013 | 0.754 | -0.041 | 0.015 | 0.007 |
| **Regional gray matter volumes** |  |  |  |  |  |  |
| Volume of thalamus | -0.007 | 0.014 | 0.616 | -0.021 | 0.016 | 0.182 |
| Volume of caudate | -0.010 | 0.014 | 0.457 | 0.009 | 0.016 | 0.568 |
| Volume of putamen | -0.052 | 0.014 | <0.001 | -0.059 | 0.017 | <0.001 |
| Volume of pallidum | 0.001 | 0.014 | 0.968 | 0.018 | 0.017 | 0.279 |
| Volume of hippocampus | -0.018 | 0.013 | 0.150 | -0.044 | 0.015 | 0.004 |
| Volume of amygdala | -0.038 | 0.012 | 0.002 | -0.070 | 0.015 | <0.001 |

### Table S8. Association between sunlight exposure time and brain structural markers after exclude dementia incidents at first ten years of follow-up. (N = 27,447) (compare to Tertile 1)

|  |  | **Tertile 2** |  |  | **Tertile 3** |  |
| --- | --- | --- | --- | --- | --- | --- |
|  | ***β*** | **SE** | **P** | ***β*** | **SE** | **P** |
| **Global measures** |  |  |  |  |  |  |
| Total brain volume | -0.025 | 0.011 | 0.026 | -0.051 | 0.013 | <0.001 |
| Volume of white matter | -0.014 | 0.012 | 0.213 | -0.032 | 0.014 | 0.021 |
| Volume of gray matter | -0.034 | 0.011 | 0.003 | -0.067 | 0.014 | <0.001 |
| Volume of white matter hyperintensities | 0.014 | 0.013 | 0.301 | 0.062 | 0.016 | <0.001 |
| **Subcortical regions** |  |  |  |  |  |  |
| Volume of thalamus | -0.029 | 0.012 | 0.015 | -0.060 | 0.014 | <0.001 |
| Volume of caudate | -0.030 | 0.013 | 0.026 | -0.040 | 0.016 | 0.012 |
| Volume of putamen | -0.022 | 0.012 | 0.065 | -0.031 | 0.014 | 0.029 |
| Volume of pallidum | -0.026 | 0.013 | 0.048 | -0.026 | 0.016 | 0.100 |
| Volume of hippocampus | -0.010 | 0.013 | 0.470 | -0.046 | 0.016 | 0.003 |
| Volume of amygdala | -0.003 | 0.014 | 0.797 | -0.031 | 0.016 | 0.052 |
| Volume of accumbens | -0.004 | 0.013 | 0.748 | -0.041 | 0.015 | 0.006 |
| **Regional gray matter volumes** |  |  |  |  |  |  |
| Volume of thalamus | -0.007 | 0.014 | 0.604 | -0.021 | 0.016 | 0.187 |
| Volume of caudate | -0.010 | 0.014 | 0.471 | 0.010 | 0.016 | 0.527 |
| Volume of putamen | -0.052 | 0.014 | <0.001 | -0.059 | 0.017 | <0.001 |
| Volume of pallidum | 0.001 | 0.014 | 0.971 | 0.019 | 0.017 | 0.261 |
| Volume of hippocampus | -0.018 | 0.013 | 0.160 | -0.044 | 0.015 | 0.004 |
| Volume of amygdala | -0.038 | 0.012 | 0.002 | -0.071 | 0.015 | <0.001 |

### Table S8. Association between sunlight exposure time and brain structural markers after excluding participants with a history of hypertension, diabetes, or stroke at baseline. (N = 20,384) (compare to Tertile 1)

|  |  | **Tertile 2** |  |  | **Tertile 3** |  |
| --- | --- | --- | --- | --- | --- | --- |
|  | ***β*** | **SE** | **P** | ***β*** | **SE** | **P** |
| **Global measures** |  |  |  |  |  |  |
| Total brain volume | -0.017 | 0.013 | 0.201 | -0.043 | 0.016 | 0.005 |
| Volume of white matter | -0.010 | 0.013 | 0.461 | -0.027 | 0.016 | 0.089 |
| Volume of gray matter | -0.023 | 0.013 | 0.087 | -0.057 | 0.016 | <0.001 |
| Volume of white matter hyperintensities | 0.025 | 0.015 | 0.105 | 0.070 | 0.018 | <0.001 |
| **Subcortical regions** |  |  |  |  |  |  |
| Volume of thalamus | -0.028 | 0.014 | 0.040 | -0.050 | 0.017 | 0.003 |
| Volume of caudate | -0.036 | 0.015 | 0.021 | -0.042 | 0.019 | 0.023 |
| Volume of putamen | -0.026 | 0.014 | 0.063 | -0.032 | 0.017 | 0.056 |
| Volume of pallidum | -0.036 | 0.015 | 0.020 | -0.029 | 0.018 | 0.110 |
| Volume of hippocampus | -0.013 | 0.015 | 0.390 | -0.035 | 0.018 | 0.054 |
| Volume of amygdala | -0.002 | 0.016 | 0.916 | -0.030 | 0.019 | 0.113 |
| Volume of accumbens | -0.007 | 0.015 | 0.630 | -0.046 | 0.018 | 0.008 |
| **Regional gray matter volumes** |  |  |  |  |  |  |
| Volume of thalamus | -0.010 | 0.016 | 0.516 | -0.021 | 0.019 | 0.266 |
| Volume of caudate | -0.005 | 0.016 | 0.774 | 0.012 | 0.019 | 0.546 |
| Volume of putamen | -0.061 | 0.016 | <0.001 | -0.050 | 0.019 | 0.010 |
| Volume of pallidum | -0.013 | 0.016 | 0.427 | 0.009 | 0.019 | 0.654 |
| Volume of hippocampus | -0.022 | 0.015 | 0.146 | -0.030 | 0.018 | 0.086 |
| Volume of amygdala | -0.031 | 0.014 | 0.029 | -0.056 | 0.017 | 0.001 |

### Table S9. Association between sunlight exposure time and cognitive tests. (N = 20,859) (compare to Tertile 1)

|  |  | **Tertile 2** |  |  | **Tertile 3** |  |
| --- | --- | --- | --- | --- | --- | --- |
|  | ***β*** | **SE** | **P** | ***β*** | **SE** | **P** |
| Numeric memory | -0.065 | 0.029 | 0.028 | -0.209 | 0.029 | 0.000 |
| Pairs matching | 0.012 | 0.013 | 0.343 | 0.011 | 0.013 | 0.385 |
| Prospective memory | 0.075 | 0.121 | 0.536 | -0.192 | 0.114 | 0.094 |
| Reaction time | 0.001 | 0.004 | 0.757 | 0.006 | 0.004 | 0.121 |
| Fluid intelligence / reasoning | -0.022 | 0.009 | 0.013 | -0.088 | 0.009 | <0.001 |
| Matrix pattern completion | -0.015 | 0.008 | 0.072 | -0.052 | 0.008 | 0.000 |
| Symbol digit substitution | -0.006 | 0.005 | 0.261 | -0.031 | 0.005 | 0.000 |
| Tower rearranging | -0.008 | 0.007 | 0.281 | -0.034 | 0.007 | 0.000 |
| Trail making 1 | 0.008 | 0.006 | 0.189 | 0.020 | 0.006 | 0.001 |
| Trail making 2 | 0.016 | 0.008 | 0.037 | 0.054 | 0.007 | 0.000 |
| Picture vocabulary | -0.163 | 0.046 | 0.000 | -0.574 | 0.045 | <0.001 |
| Paired associate learning | -0.010 | 0.009 | 0.275 | -0.052 | 0.009 | 0.000 |

**Figure S1.** The restricted cubic spline of natural sunlight exposure with the volumes of subcortical regions and gray matter regions

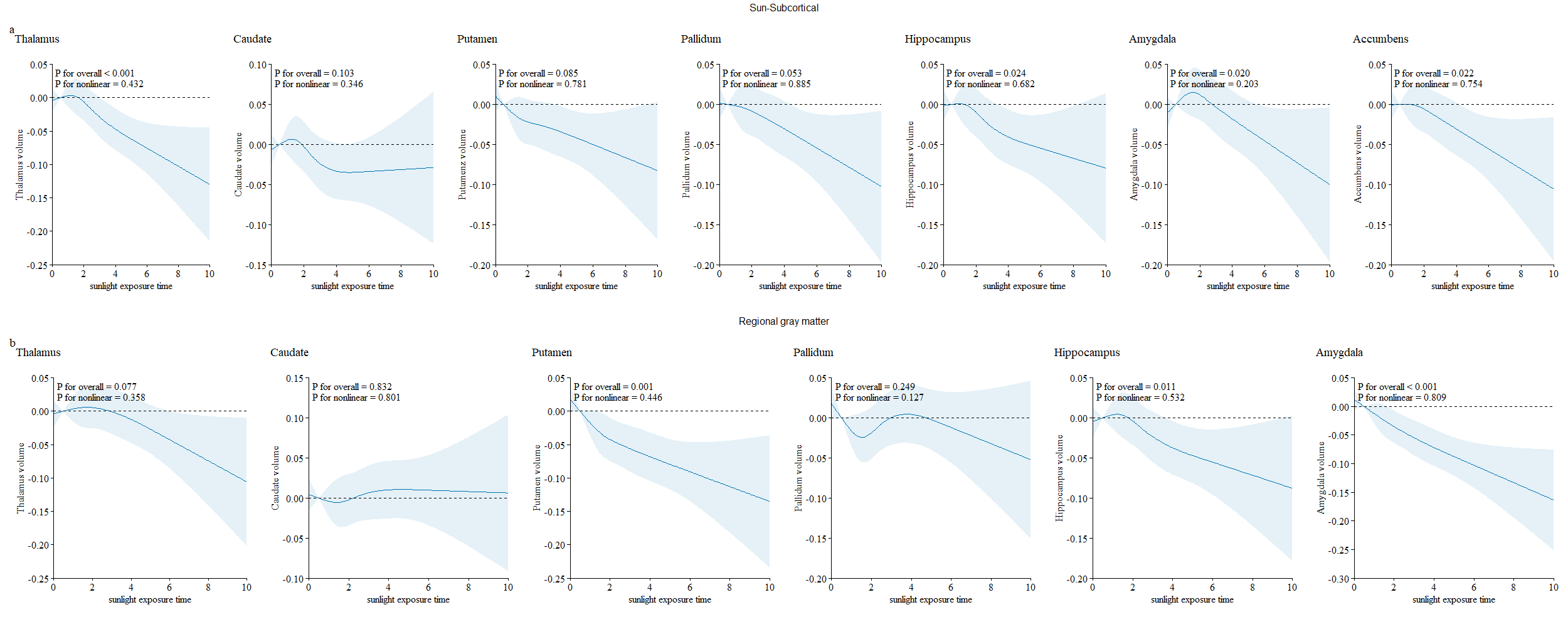

a: the volumes of subcortical regions

b: the volumes of gray matter regions

**Figure S2.** The restricted cubic spline of natural sunlight exposure with the volumes of subcortical regions and gray matter regions stratified by season.

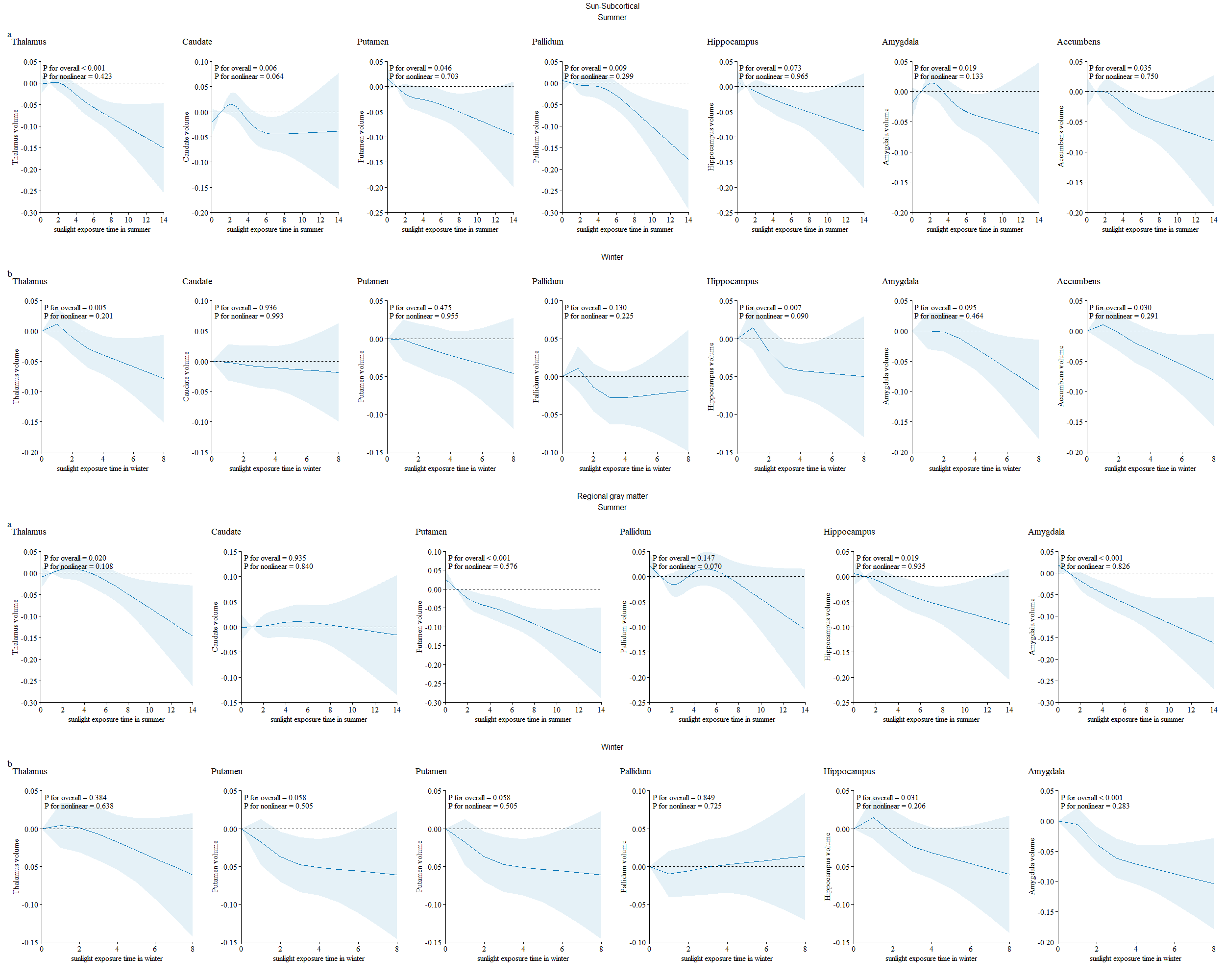

a: natural sunlight exposure in summer.

b: natural sunlight exposure in winter.
